# DuckDerm: An Improved Water Infiltration Prevention Product for Long-Term Intravenous Therapy Patients

**DOI:** 10.1101/2024.11.25.24317953

**Authors:** Dalia Fantini, Abigail Fisk, Andres Leon-Vargas, Emma Lynch, Christine Scarlett, Devon Stafford, Marcos Zegarra, Brady John Bielewicz

## Abstract

We propose an alternative to the widely implemented intravenous (IV) shower guard solution, AnchorDry, which we have coined DuckDerm. The AnchorDry is a rigid square of plastic with an adhesive border on the outer edge. This design does not adhere to the arm during the bathing period as it cannot withstand the range of motion (ROM) patients exert, leaving the Tegaderm and IV site vulnerable to water intrusion. Significant water intrusion compromises the IV site, leaving it susceptible to infection. By interviewing seasoned clinicians, nursing faculty, students, and patient care technicians (PCTs) we ascertained ineffective shower guard fit is an issue rooted in design and material selections. We have found that nurses and PCTs are often getting creative to modify or make their own IV shower guards.

After several design iterations and consultations with several University of Pittsburgh nursing faculty, we mass-manufactured our design. The DuckDerm body is composed of 1 mil polyethylene plastic, which is biocompatible and able to move with the skin, a hypoallergenic adhesive, and a foam barrier to envelop the Tegaderm. Based on this initial testing, we formulated our design specifications: (1) maintaining the patient’s ROM, (2) ease of use for both patients and clinicians, and (3) preventing water infiltration. We provided our manufactured design and interviewed nursing students and faculty (n=21) to address the following criteria: (1) the DuckDerm must be simple for clinicians to apply to not take away time from more essential tasks and to encourage patient self-application; (2) the DuckDerm must retain functionality without providing discomfort to the patient during use and removal. We also conducted functionality testing using our manufactured design to address the following criteria on volunteers (n=11): the DuckDerm must (1) contain all the IV components within the foam barrier, (2) not place unnecessary pressure on components to dislodge, and (3) fit securely on the patient’s arm. Additionally, the Duckderm must adhere to the skin while subjected to running water and movement, and the DuckDerm must be able to provide a sufficient range of motion about the antecubital space to allow for arm movements required while showering. The Duckderm outperformed the currently utilized solution in both verbal feedback and assessment by potential users (nurses and PCTs) and in functionality testing; therefore, we propose our novel design as an alternative to the currently employed AnchorDry.

## Introduction

### Surmising the Clinical Need

Approximately 80% of hospitalized patients in the United States require an intravenous (IV) line to deliver fluid, medicine, or blood [1]. When taking a shower, patients are required to use a waterproof wrapping (what we call in this work a shower guard) to secure the line and prevent the infiltration of water and soap to the IV insertion site. The shower guard does not directly interface with the IV site but functions to protect the Tegaderm wrapping.

Nosocomial infections, also known as healthcare-associated infections (HAIs), frequently result from the use of intravenous (IV) devices, such as central venous catheters (CVCs). These infections are primarily caused by microbial colonization of the catheter’s surface or contamination during insertion, leading to bloodstream infections. Proper aseptic techniques, timely removal of unnecessary catheters, and the use of antimicrobial-coated devices can reduce infection rates. Nosocomial bloodstream infections are associated with increased morbidity, mortality, and healthcare costs, emphasizing the need for stringent infection control measures [2,3]. Nosocomial infections related to intravenous (IV) devices can be exacerbated by inadequate protection of the IV site during a patient’s hospital stay, particularly during activities such as showering. Moisture exposure can compromise dressings, creating a pathway for microbial entry and increasing the risk of bloodstream infections. To mitigate this risk, protective measures such as waterproof dressings, specifically designed catheter covers, and proper patient education are critical. Regular inspection of the IV site for signs of moisture, damage, or infection further enhances safety. Adherence to these strategies is essential to maintaining the integrity of the IV site and reducing infection risks [4, 5].

Current water-resistant shower guards are restrictive (**Fig 1A** and **B**), limiting patient mobility, and are typically ill-fitting. In addition, the current solution fails early and often during the bathing period, requiring clinicians to fashion their own shower guard alternative. After stumbling upon this clear clinical need, we propose our redesigned shower guard (the DuckDerm, **Fig 2A-C**). This design seeks to prevent water infiltration and allow for an increased range of motion (ROM), while remaining adhered to the arm for the duration of the bathing period.

**Figure 1.**
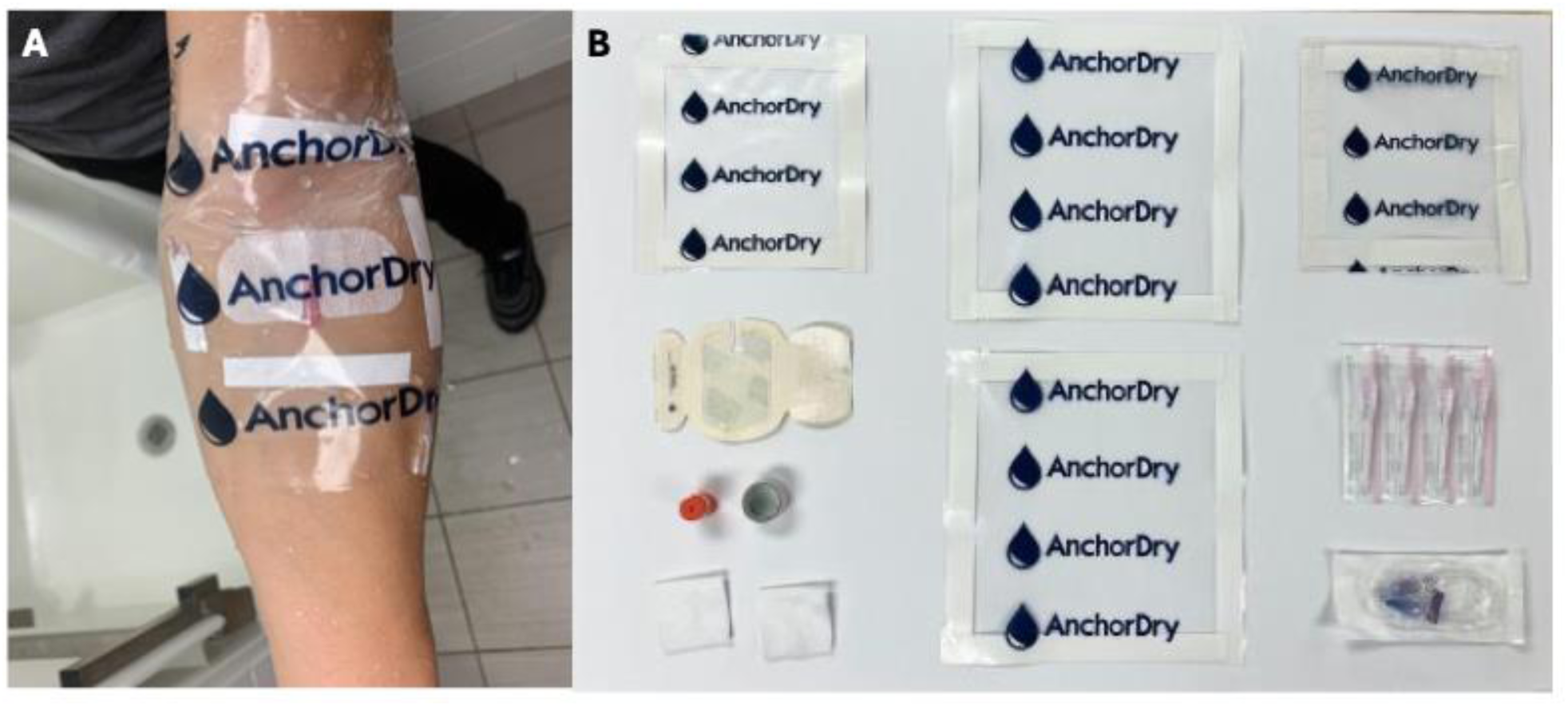
(A) Image shows the currently implemented clinical solution (AnchorDry) overtop the mock intravenous site, complete with a Tegaderm; (B) Image shows an IV access kit, complete with an IV catheter, TegaDerm dressing, an IV extension set, alcohol and gauze prep pads, and Transpore tape.

**Figure 2.**
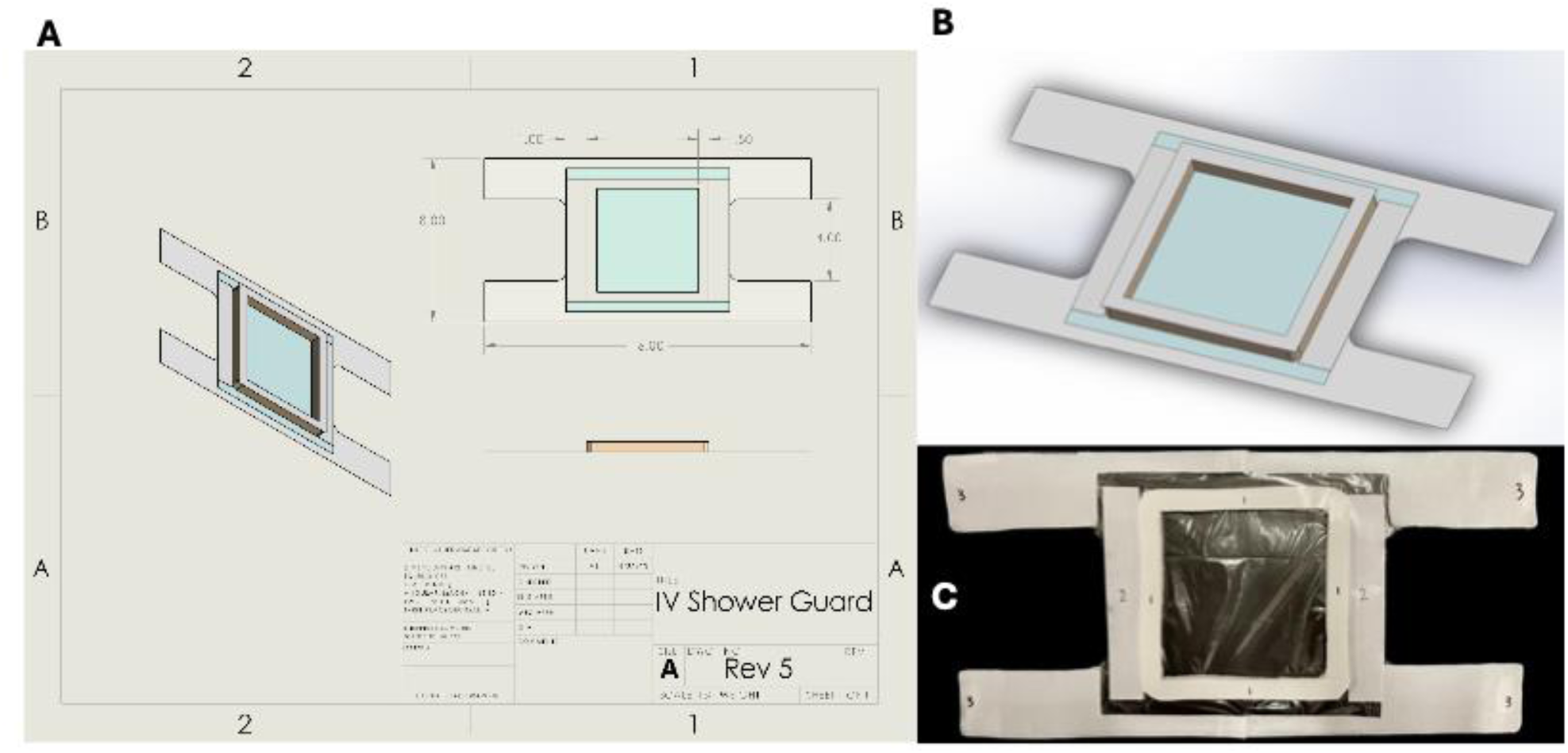
A SolidWorks (A, B) and physical representation (C) of the most updated revision following design changes based on qualification testing.

## Results

### Final Prototype

We have identified the most prevalent hazards and associated risks for the shower guard design: 1) water enters the IV insertion site, 2) IV dislodgement, 3) skin irritation when removing the shower guard, and 4) bacterial contamination of the shower guard (summarized in **Table S6** and **S7**). The final iteration of the DuckDerm is showcased in **Fig 2C**.

The proposed shower guard device is designed with specific dimensional, physical, and material specifications to ensure functionality, comfort, and safety. The straps on the top and bottom measure 16 inches in length and 2 inches in height but can be trimmed to fit various arm circumferences. The foam component, with a thickness of 0.5 inches, covers a 6×6 inch square to effectively conceal the IV site, Tegaderm dressing, and extension tubing. A waterproof adhesive lines the edges of the guard and foam layer, providing secure attachment, as shown in Figure 1. Dimensional tolerances are determined by the 0.25-inch width of the marker used to trace the plastic template, and the product’s final mass remains to be determined to ensure it does not impede patient movement.

The general size requirements include a strap height of 2 inches ± 0.25 inches, a strap length of 16 inches ± 0.25 inches, a plastic body height of 7 inches ± 0.25 inches, and a plastic body length of 8 inches ± 0.25 inches. Labels will clearly mark the proximal and distal ends, alongside numbered peelable adhesive covers indicating the order of application. A set of written instructions accompanies each unit, providing step-by-step guidance on proper usage.

Designed for single use, the shower guard is disposable and does not require cleaning after use. Its packaging ensures sterility and protection from contamination until application. Materials can withstand water temperatures ranging from 36°C to 65°C and humidity levels associated with hot showers. The guard’s construction includes high-tensile-strength polyethylene sheets, skin-safe medical-grade adhesive, and absorbent foam to prevent water from reaching the IV insertion site. All materials are hypoallergenic, water-resistant, and cost-effective, making the product accessible for short-term use in medical settings.

Finally, the device must be stored in a clean environment alongside other disposable medical supplies. It is not intended to interface directly with wounds, negating the need for sterilization. However, its design allows for contact with alcohol-based solutions, commonly used for hospital sterilization, ensuring compatibility with routine medical practices.

### Validation Testing

#### Shower Guard Assessment

Visualization of the data collected in the shower guard assessment by clinical users is summarized in **Fig 3A**, and time for application by these users, correlated to patient care experience (**Fig 3B**). The survey’s first question asked whether participants believed the product would remain securely attached to the antecubital region during typical showering motions. The average response was 8.14, categorized as passive.

**Figure 3.**
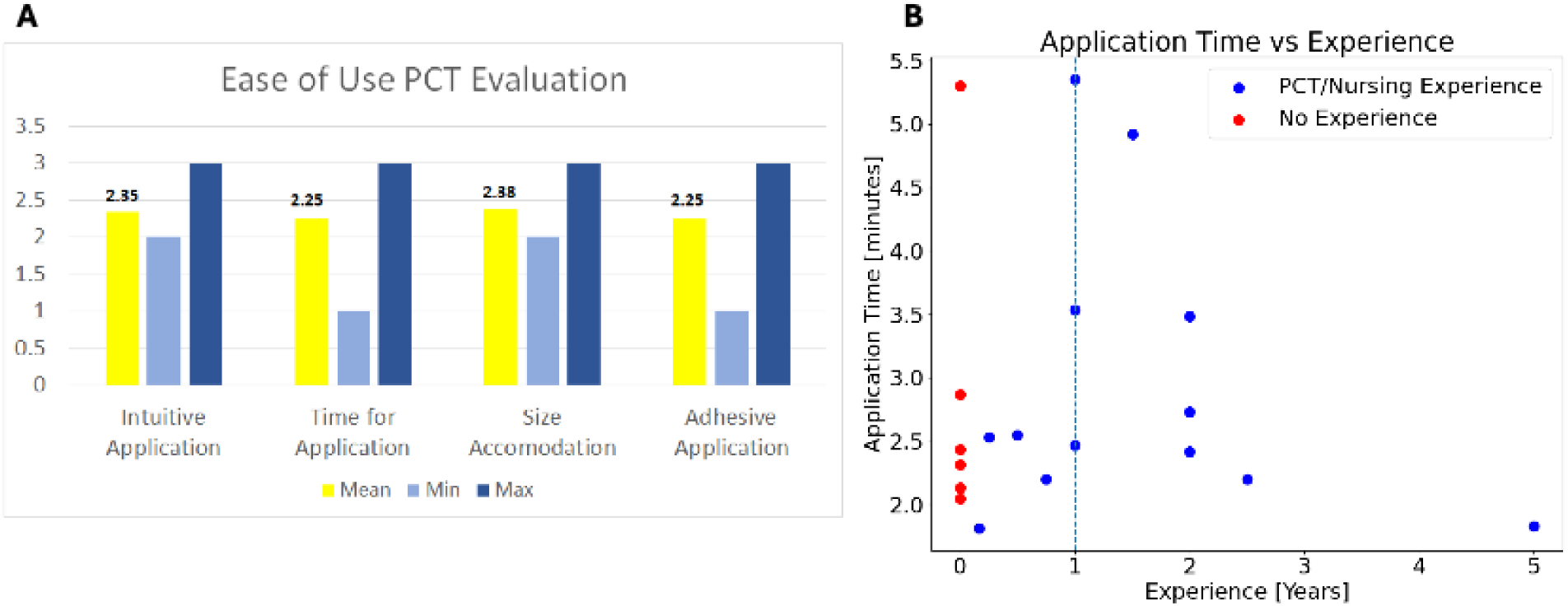
(A) Graphical representation of responses on a scale of 0 to 10, outlining users responses for intuitive application, time for application, size accommodation, and adhesive application; (B) Plot outlining application time for the DuckDerm versus duration of clinical experience.

Question 2 assessed whether the foam barrier would protect the IV site from water intrusion if the surrounding plastic adhesive detached, yielding an average score of 7.82 (passive). Question 3 asked if a primary caretaker (PCT) could easily and quickly apply the product, scoring 7.86 (passive). Question 4 evaluated the clarity and intuitiveness of the application instructions, scoring 8.50 (passive). Question 5 asked if the product appeared more effective than commonly used alternatives, such as AnchorDry, and received a promoter score of 9.14. Question 6 evaluated the product’s comfort during arm movement, with an average score of 8.45 (passive). Finally, Question 7 assessed whether the adjustable straps made the product suitable for any patient, scoring 7.82 (passive). Overall, the survey’s average score was 8.24 out of 10, which falls within the passive range but exceeds the acceptance threshold of 65%. Therefore, DuckDerm successfully passed this evaluation.

#### Ease of Use Testing

Ease of use rating, experience, and application time were recorded (experimental set-up is highlighted in **Fig 4A**), with the distribution of those ratings displayed in **Fig 4B** and **4C**. We found that well over 75% of participants found the product to be satisfactory or higher in each ease-of-use category. 65% of participants found the shower-guard satisfactory in the category of intuitive application, size accommodation, and adhesive application. Those same categories received exceptional ratings in 35%, 35%, and 30% of responses respectively. Time for application had 55% of participants rate it satisfactory and 35% exceptional ratings. This gave a combined response of satisfactory or greater of 100%, 90%, 100%, and 95% in intuitive application, time for application, size accommodation, and adhesive application respectively. The average of the satisfaction score is shown in figure 2. We wanted to check the time for application relative to clinical experience for potential trends and found that time for applications decreases with years of experience after a one-year minimum experience (Figure 3).

**Figure 4.**
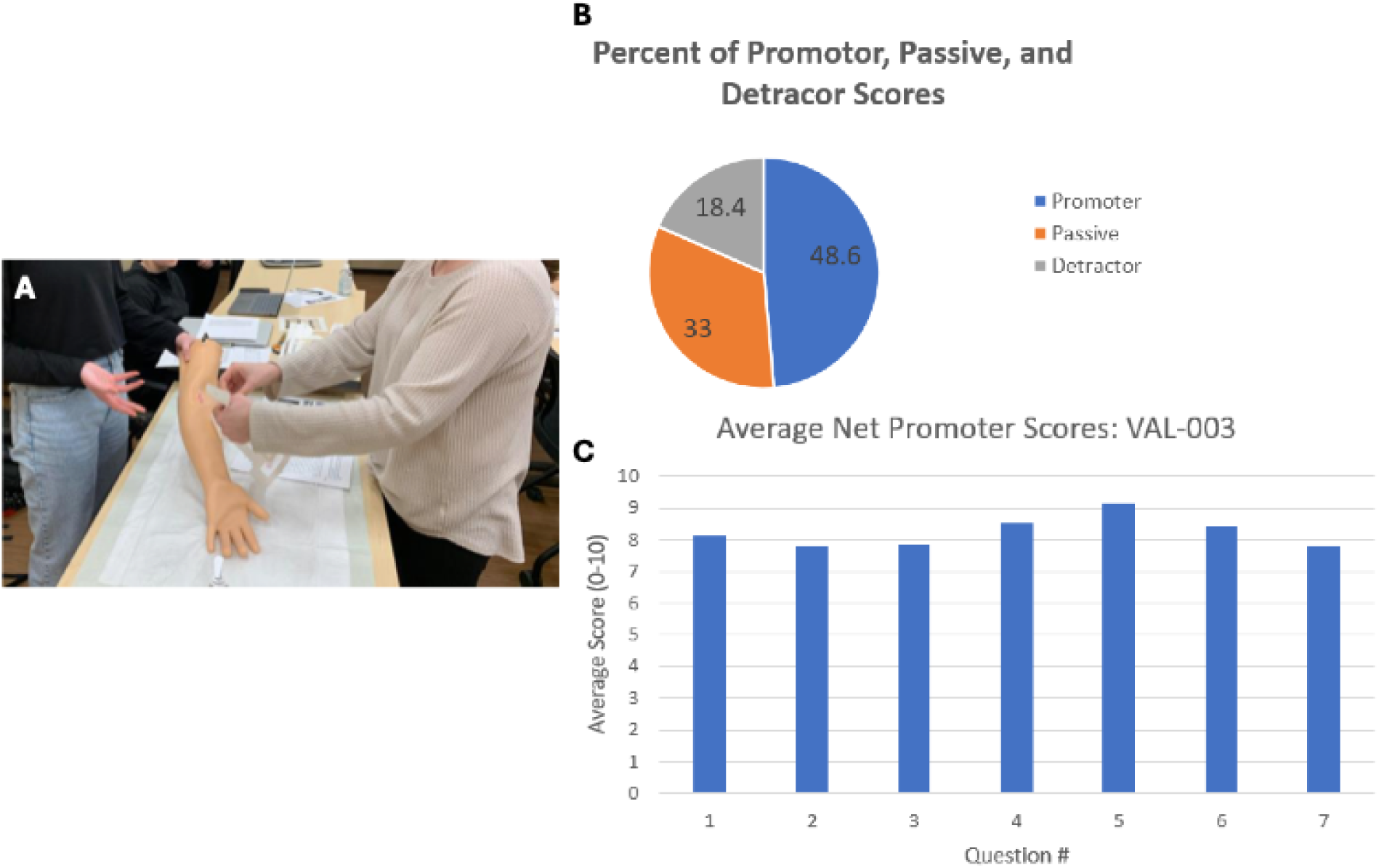
(A) Test set-up, including a facilitator, note taker, participant, instructions sheet, mannequin arm with simulated IV, and shower guard prototype; (B) Average percent of responses to all questions given in the promotor, passive, and detractor categories; (C) Graphical representation of the average scores on questions posed to potential clinical users.

#### Verification Testing Comfortability Testing

The comfort and pain ratings are summarized in Figure 5A, revealing generally low scores: average comfort during wear was 1.73, average pain during wear was 1.00, and average pain during removal was 2.73. These results indicate that participants found the product comfortable to wear, with minimal pain reported during wear or removal. All scores were below the failing threshold, confirming the product passed the comfort assessment. Wear duration did not significantly affect comfort or pain scores (Figures 5B and 5C), as participants who wore the product for extended periods reported similar levels of comfort and pain as those who wore it for shorter durations.

**Figure 5.**
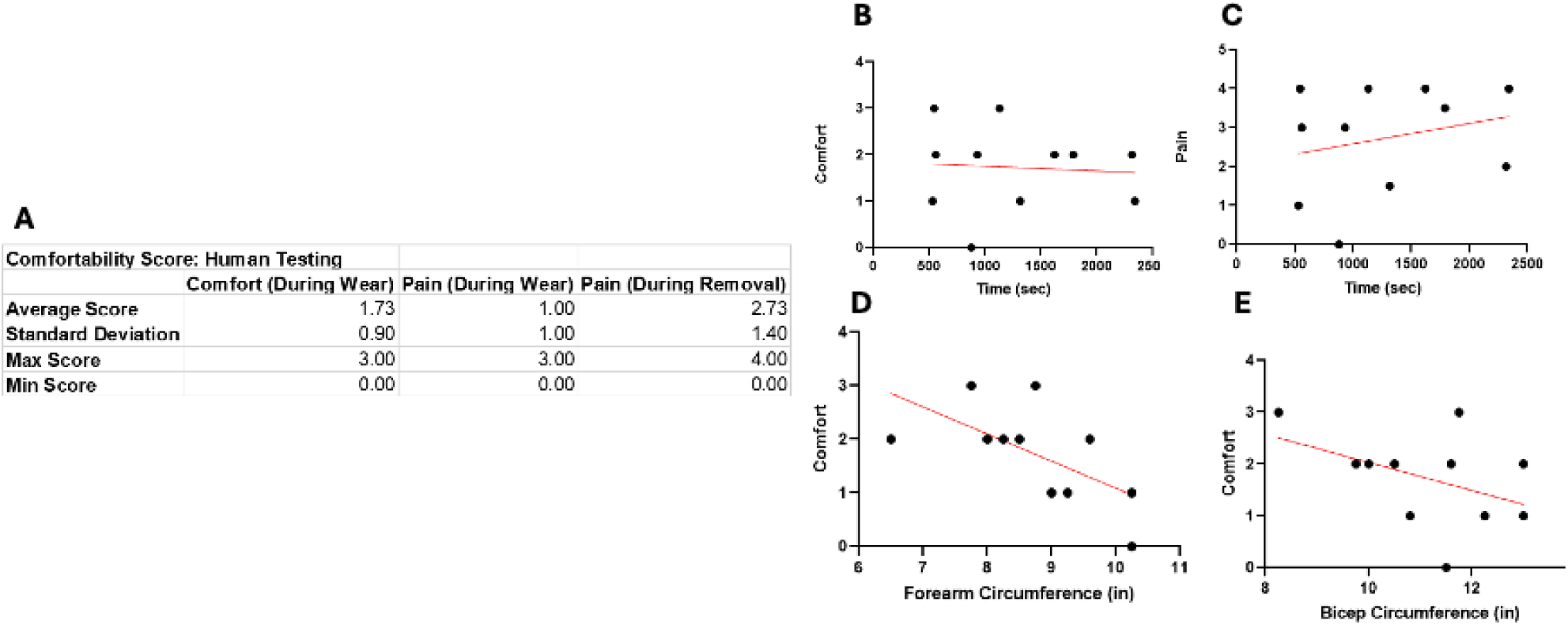
Overall comfortability scoring (A), comfort and pain scores correlated to time (B, C), and comfortability scoring correlated to forearm circumference and bicep circumference (E).

While bicep circumference showed no significant correlation with comfort scores (Figure 5E), forearm circumference was slightly negatively correlated with comfort scores (r = - 0.623, p = 0.041; Figure 5D). This suggests that participants with smaller forearm circumferences found the product less comfortable compared to those with larger forearms.

#### Range of Motion Testing

The DuckDerm must be able to provide a sufficient range of motion about the antecubital space to allow for arm movements required while showering. We assessed elbow flexion, extension, and rotation about the elbow as well as pronation and supination of the wrist to simulate movements performed during a shower. Each motion was assigned a score of 0, 1, or 2, correlating to complete fail, minor fail or pass, respectively. A score greater than or equal to 11 out of 20 possible points was considered an overall pass whereas anything below that was considered a fail. Out of the eleven participants tested, ten of them received an overall passing score (**Table 1**). The average score for all the participants was 18.5, indicating the DuckDerm did not inhibit or limit participant mobility by our predetermined threshold score of 15.

**Table 1.** Range of Motion score for each participant correlated to an overall pass or fail for the protocol, accompanied by general notes logged by the tester from each session. Bicep and Forearm circumference were also recorded for each subject.

| Test Subject ID | Range of Motion Score | Bicep Circumference (in) | Forearm Circumference (in) | Pass | Fail | General Notes |
| --- | --- | --- | --- | --- | --- | --- |
| H01 | 17 | 13 | 10.5 | X |  | Tighter fit at the proximal end. Peeling by the straps. |
| H02 | 20 | 9.75 | 8 | X |  | Catheter/extension tubing limited flexion/extension. Minor peeling at proximal end. |
| H03 | 20 | 10 | 6.5 | X |  | Good ROM. No peeling. |
| H04 | 20 | 10.5 | 8.25 | X |  | Catheter/extension tubing limited flexion/extension. |
| H05 | 20 | 12.25 | 9.25 | X |  | No peeling. Catheter/extension tubing poked skin. |
| H06 | 18 | 11.75 | 8.75 | X |  | Tight around the straps. Peeling by proximal/distal edges of straps. Catheter/extension tubing poked skin. |
| H07 | 20 | 10.8 | 9 | X |  | No limitations in range of motion. |
| H08 | 20 | 11.6 | 8.5 | X |  | Adhesive pulled on hair, not skin. |
| H09 | 10 | 13 | 9.5 |  | X | Peeling at proximal and distal ends of the straps. |
| H10 | 20 | 11.5 | 10.25 | X |  | N/A |
| H11 | 18 | 8.25 | 7.75 | X |  | Minor limitation on proximal end from bicep. |

#### Shower Simulation

The DuckDerm must adhere to the skin while subjected to running water and movement (**Fig 6A** and **Fig 6B**). This test examines the functionality of the shower guard over an allotted 5 minutes while the participant performs flexion/extension, pronation/supination, and rotation. This test primarily determines the efficacy of the primary and secondary barriers implemented in the design, the adhesive, and the foam, respectively. Each participant placed their arms directly into the stream of water while performing the aforementioned movements. Water indicator tape lined the IV and extension tubing along the antecubital space. Upon contact with water, the tape would turn a bright red color. Activation of the water contact tape would result in a “fail”, with the time of failure recorded. Of the 8 participants who have undergone this testing, 4 received a passing score (**Fig 6C**). Of the 4 participants who received a “fail” on this test, the average time of failure is 125.75 seconds (or 2 minutes and 5.75 seconds). Minimal correlation has been detected between bicep circumference (**Fig 6D**) or forearm circumference (**Fig 6E**) in regard to DuckDerm adherence.

**Figure 6.**
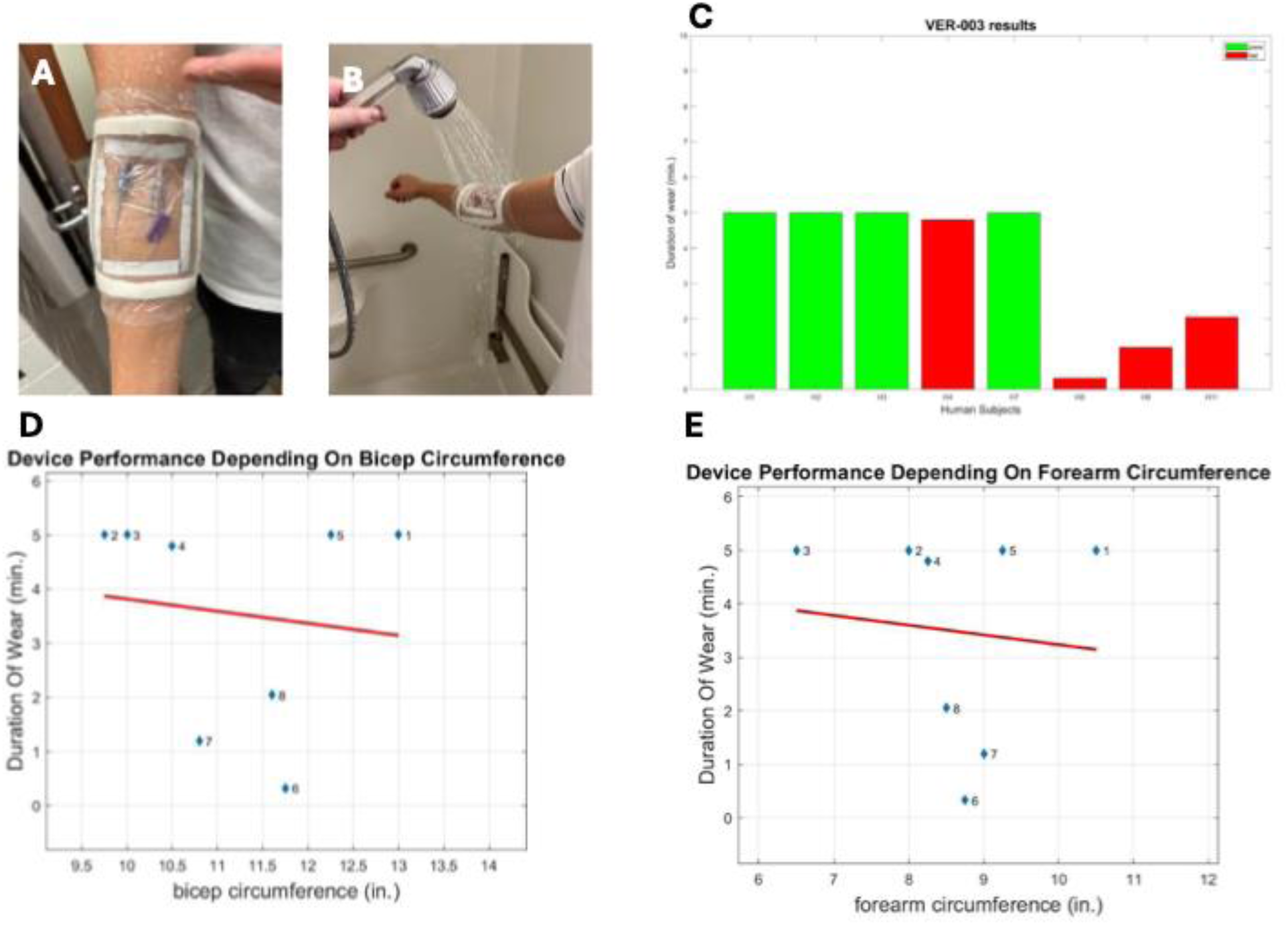
(A) Image shows correct product placement of the DuckDerm; (B) Experimental set-up for the shower simulation testing; (C) A box plot representation of the human subjects who passed during the entire duration of the protocol (which will be 5 min.) and those subjects who failed and at what point in time they failed; (D) A scatter plot of the bicep circumference taken from the subjects and their duration of wear, with the red line being the best fit line of the plot; (E) A scatter plot of the forearm circumference taken from the subjects and their duration of wear, with the red line being the best fit line of the plot.

## Discussion

The development of DuckDerm began by gathering valuable insights from relevant stakeholders, including healthcare providers and patients, who were interviewed about their experiences with existing waterproof IV shower guards like the Shower Shield and AnchorDry. These products highlighted key challenges that needed addressing, such as ensuring the guard stayed securely adhered to the arm during movement, preventing water exposure to the IV site, ease of application by caretakers, and comfort for extended wear during a shower. The ethnographic data collected formed the basis for identifying critical requirements for DuckDerm.

To meet these needs, we engaged with plastics and adhesive manufacturers, along with professionals in relevant fields, to determine the material requirements for the product. Our final design incorporated 1 mil polyethylene plastic for the body of the DuckDerm. This material is biocompatible and flexible, allowing it to move with the skin, reducing discomfort during use. We also chose a hypoallergenic adhesive to ensure that the product adhered securely to the skin without causing irritation. Low-resolution iterative prototypes were created to test different geometries, ensuring the design functioned effectively before advancing to formal testing.

The validation and verification testing of DuckDerm, as outlined below, demonstrated that the product met its design requirements. These testing phases supported our choice of materials and design features, confirming the potential of DuckDerm to address the significant gap in the market for a reliable, comfortable, and effective IV shower guard. Given that approximately 80% of hospitalized patients require an IV line at some point [1], DuckDerm could provide a much-needed solution to enhance patient care, prevent complications, and improve the overall patient experience during hospitalization. This thorough, stakeholder-driven approach has resulted in a product that not only solves practical issues but also ensures comfort and security for the patient, representing a promising innovation in patient care.

## Methods

### Initial Prototyping

In the first revision prototype (**Fig S1 A**) our group produced, we were eager to implement a crisscrossing arm band component to ensure proper fit to the IV site. The material utilized in this iteration was 6 mil polyethylene (PE) plastic, velcro, and gauze attached to the PE by glue as the absorptive element. The velcro was used for prototype handling and ease of use testing. We had determined the plastic thickness used in this design was far too thick, constricting the patient’s range of motion and was uncomfortable during use. A critique from our project mentor was the ease of use of the shower guard. With the long straps, it would be difficult for the clinician to put on the shower guard, let alone for the patient to do it independently. Additionally, the gauze was found to be far too small to envelop the Tegaderm and prevent water infiltration.

In the second revision (**Fig S1 B**), we acquired a more suitable plastic for our prototype, shifting to 1 mil polyethylene. We found that while this plastic was more difficult to work with when placed on a mannequin or one of our arms, the plastic film was better able to flex with movement. Another notable revision was the shortening of the plastic straps to cut down on plastic usage for usability and sustainability purposes. A double-sided adhesive was also implemented to determine the ease of application.

The gauze as the absorptive component was kept from the previous revision; however, we were initiating contact with materials experts and companies to find medical-grade materials to incorporate into the final design.

For the third revision (**Fig S1 C**), we sought to increase the surface area of the adhesive placement following a conversation with our project mentor. Additionally, we transitioned from the gauze present in the first two iterations following a series of killer testing experiments to determine the water-absorbing capacity of the Ca-alginate gauze versus wound care foam. The wound care foam significantly outperformed the gauze, and we pivoted to implement foam in our design. Additionally, we opted to place a double-sided adhesive film onto the foam itself to provide an additional water barrier.

### DuckDerm Manufacturing

Laser-cut templates were developed for the DuckDerm in accordance with final design specifications to enable mass production for validation and verification testing. These templates ensured precise cutting of the medical-grade foam and plastic film. The adhesives utilized included Waterproof Transparent Film Roll (Housables), 3M Super 77 spray adhesive, and Vapon double-sided tape. The final mass-manufactured design incorporated medical-grade foam (MedPride Foam Dressing) and a 1-mil medical-grade polyethylene film.

### Validation Testing

Once our protocols were vetted and approved by the University of Pittsburgh internal review board, we were able to recruit a total of 22 participants, all of whom are either current or student nurses, patient care technicians, and University of Pittsburgh nursing faculty members to participate in our validation studies.

### Shower Guard Assessment

Participants begin by reading over the provided application instructions (refer to (**Fig S2**) for the product, and then are asked to apply the product to a mannequin arm with an IV site simulated in the antecubital region. After the subject has applied the shower guard, they are asked for their feedback on the product. The participant is then asked about the ease of use, design, comfort, effectiveness, and sizing of the product. Seven questions are asked on a scale from zero to ten, zero being strongly disagree with the statement, and ten being strongly agreeing with the statement. Seven statements were posed of the subject in the survey for this study and are as follows:

1. This product will remain adhered to the antecubital region through the typical range of motion seen while showering.
2. A primary caretaker will be able to easily and quickly apply this product.
3. The foam barrier will protect the IV site from water intrusion if the surrounding plastic adhesive detach from the arm.
4. The instructions to apply the product are clear and intuitive.
5. The product appears comfortable for the patient to wear and move their arm in.
6. The adjustable length of the straps will allow this product to fit on any patient.
7. This product appears more effective than the current product used (AnchorDry). These questions were all ranked on a scale from 0 (strongly disagree) to 10 (strongly agree). A net promoter score was given, with a score from 0 to 6 as a detractor, 7 and 8 being passive, and 9 and 10 being a promoter. Twenty-two responses were collected when this test was administered.

### Ease of Use Assessment

To obtain feedback from PCTs and nurses on the ease of use of the shower guard, we provided subjects with the novel shower guard and a mannequin arm with artificial skin for use in clinical simulations. We also provided two versions of application instructions that would be included in a market-distributed version of the shower guard. Participants had an opportunity to review the instruction sheet they felt was most intuitive to use.

Once satisfied with reviewing the instructions, participants were timed as they began applying the shower-guard on the mannequin arm with no feedback from the research team. The no-feedback policy was used to properly gauge the intuitiveness of the application in a market-setting, where the development team would be unable to provide feedback. Upon completion of the application, the time of application was recorded. We then asked for feedback from the participants on the relative ease of use of the product in comparison to existing shower-guard solutions. We asked participants to rate the ease of use in terms of intuitive application, time for application, and sizing accommodations. Intuitive application refers to the ability to apply the shower-guard with minimal or no feedback in a manner that the individual applying it is confident and satisfied with the final application. Time for application refers to their satisfaction with the time taken to apply, it is not too long and becomes cumbersome in a clinical setting. Size accommodation refers to the ability to apply the shower guard, with or without rapid modifications (trimming of arm bands), on varying arm sizes. Adhesive application refers to the ease of working with the adhesive used on the shower-guard, usually referring to ability to ensure the adhesive attaches to desired locations with minimal effort and preventing unwanted self-adhesion. These were all rated on a scale of 1 to 3, with 1 being unsatisfactory, 2 satisfactory, and 3 extremely satisfactory. Experience of participants in nursing or PCT was also recorded.

### Verification Testing

Once our protocols were vetted and approved by the University of Pittsburgh internal review board, we were able to recruit participants from the ages of 20 to 29 with 4 males and 7 females (n=11 participants) to participate in our verification studies.

### Comfortability

Comfortability was assessed at the end of human testing sessions following successful application of the DuckDerm (**Table S1**). Time of wear was taken as soon as the product has been removed. The participant was then be asked if the product was comfortable to wear. Verbal feedback was recorded. The participant was then asked to score their comfort during wear using the provided ranking scale. A lower score correlates with the product being more comfortable. The participant was then asked if the product caused them any pain during wear. Verbal feedback was recorded. Using the provided ranking scale, the participant can be asked to provide a score for their pain during wear. A lower score correlates with less pain. Finally, the participant was then be asked if the product caused them any pain during removal. Verbal feedback was recorded. Using the provided ranking scale, the participant was then asked to provide a score for their pain during removal. A lower score correlates with less pain.

### Range of Motion Testing

Prior to beginning the test, anthropometric measurements were taken for each subject, specifically biceps and forearm circumference. The DuckDerm was placed on the test subject’s antecubital space, according to the supplied directions (**Fig S2**), over the IV simulation materials. The subject was then asked to stand in anatomical position to begin the first set of range of motion exercises, starting with 3 to 4 rounds of flexion and extension at the elbow. The subject then adjusted their body so that their arms were straight out in front of them creating a 90° angle with the front of the trunk of the body while the arms remain perpendicular to the chest. From this position, the subject was asked to perform 3 to 4 sets of pronation and supination about the wrist followed by 3 to 4 sets of flexion and extension about the elbow. From this same position with the arm flexed at the elbow, they rotated their forearm about the elbow 3 to 4 times. The subject then transitioned to the final body position which was a T-pose, where the extended arms created a 90° angle with the side of the trunk of their body. They again performed 3 to 4 sets of flexion and extension about the elbow moving their arm towards the head, followed by another 3 to 4 rotations about the elbow to simulate scrubbing their scalp in the shower. After each of these movements, the DuckDerm was analyzed to assess whether there was any peeling of the main plastic body from the subject’s arm. A score was assigned to the subject for each of the movements depending on the amount of peeling or movement inhibition that occurred.

### Shower Simulation

Moisture indicator tapes were placed surrounding the IV components, with the DuckDerm enveloping both the Tegaderm and indicator tapes. With these in place, the subject was instructed to place their arm in the shower under running water. While the participant had their arm under a stream of water, the facilitator of the test instructed the subject to perform a set of arm movements. For each movement instructed by the facilitator, the subject performed that movement for 30 seconds. The facilitator recorded any relevant notes or feedback about the device.

## Ethics Declaration

We ascertain that all interviews, validation testing, and verification testing were done in collaboration with and approved by the University of Pittsburgh Internal Review Board (IRB) and have acquired exempt status by the IRB. All data has been deidentified and cannot be linked to individual participants.

## Data Availability

All data produced in the present study are available upon reasonable request to the authors

## Acknowledgements

The authors would like to thank Dr. Mark Gartner, our Bioengineering faculty advisor, for his advice and guidance during the design and validation and verification testing process.

## Supplemental Information

**Figure S1.**
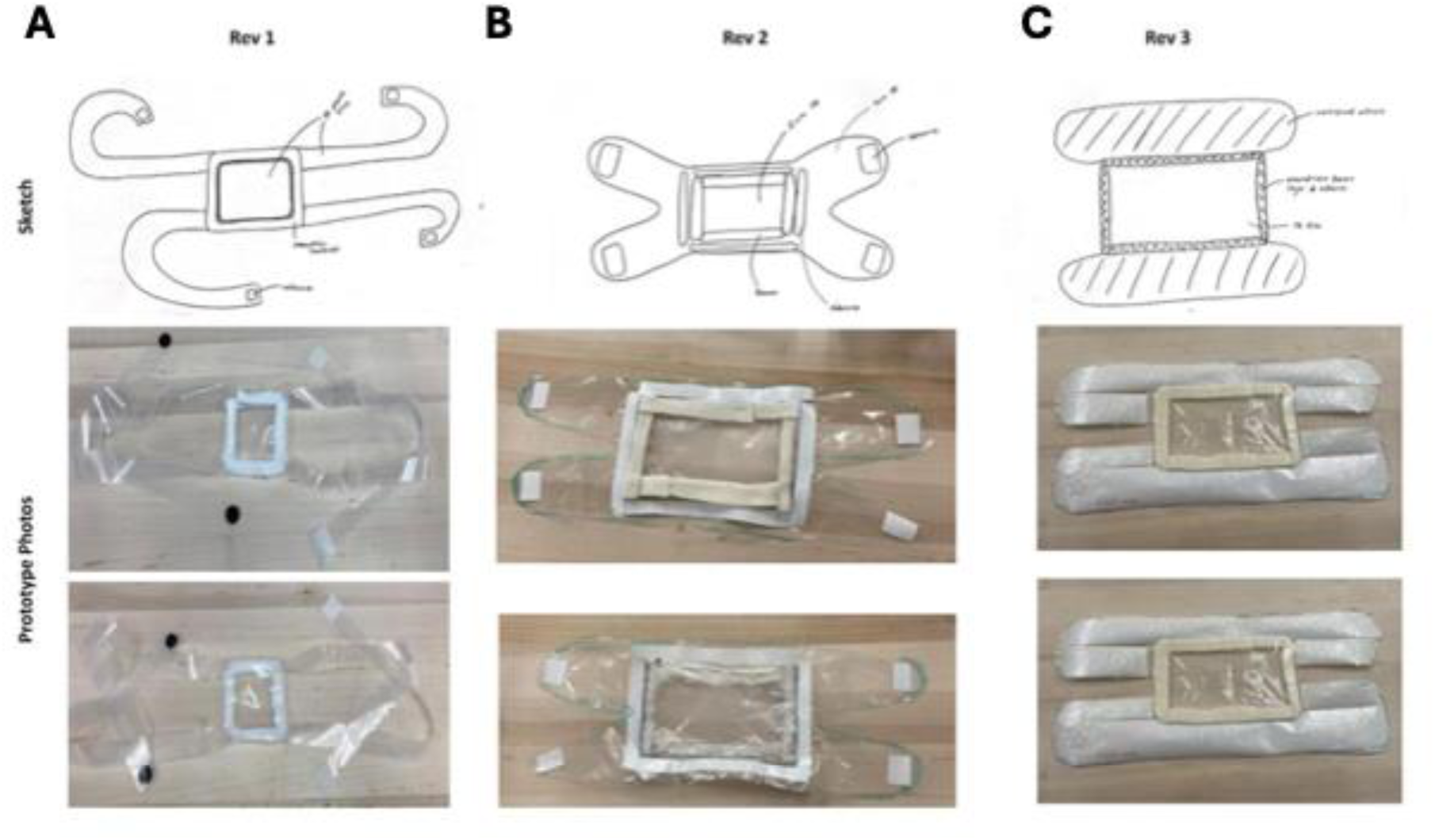
Images and sketches above recapitulate the design evolution of our prototype over the series of three significant revisions (A-C).

**Figure S2.**
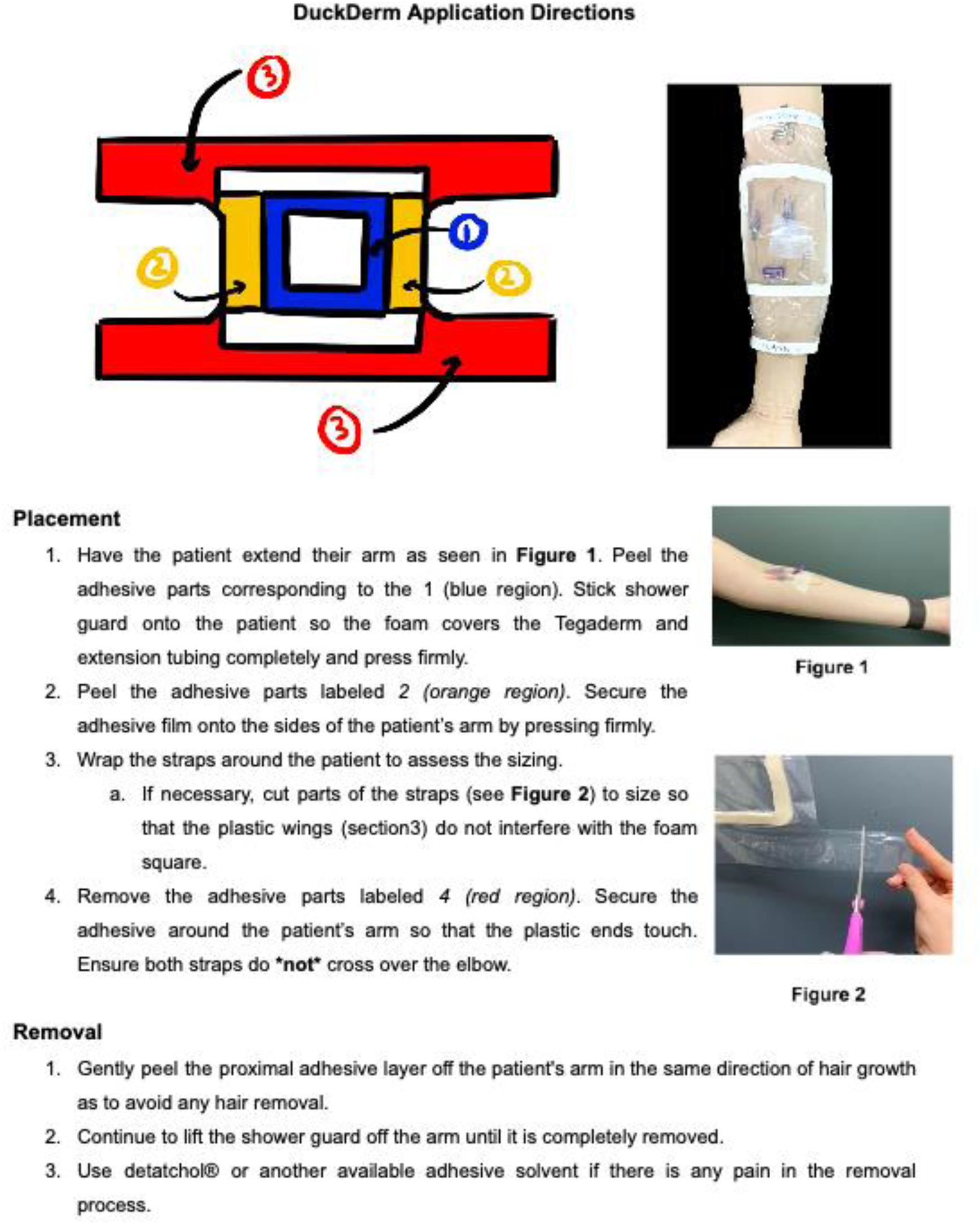
Directions for the placement and removal of the DuckDerm complete with a color-coded schematic and photos showing correct placement and an example of customization for patient fit.

**Table S1.** Passing (✔) or failing (✖) of test criteria accompanied by general notes logged by the tester from each session.

| Test Subject ID | Secure Fit Around Arm | Contains All Components | No Dislodging of Components | Overall | Notes |
| --- | --- | --- | --- | --- | --- |
| H01 | ✓ | ✓ | ✓ | ✓ | Only extension tubing was utilized for this testing, no catheter hub and Tegaderm was too small to fully cover the extension tubing. Adhesive is thin at areas and the adhesive strips were cut for fit |
| H02 | ✓ | ✓ | ✓ | ✓ | The purple component on the extension loop is up against the plastic. Straps were cut, still overlap around back of arm |
| H03 | ✓ | ✓ | ✓ | ✓ | Straps were not cut and overlap with the foam barrier section, distal part rolled up slightly and lifts up from the arm |
| H04 | ✓ | ✓ | ✓ | ✓ | Parts of the IV are pressing against the plastic |
| H05 | ✓ | ✓ | ✓ | ✓ | N/A |
| H06 | ✓ | ✓ | ✓ | ✓ | N/A |
| H07 | ✓ | ✓ | ✓ | ✓ | N/A |
| H08 | ✓ | ✓ | ✓ | ✓ | N/A |
| H09 | ✓ | ✓ | ✓ | ✓ | Pressure on the IV components which may cause problems with the plastic. Some adhesive seemed to not be fully secure. Could be due to body hair or weak adhesive |
| H10 | ✓ | ✓ | ✓ | ✓ | Application of top strap covers elbow minimally. Slight adhesive issue at top |
| H11 | ✓ | ✓ | ✓ | ✓ | N/A |

**Tables S2-5:** Tables outlining the severity scale (Table S2), probability scale (Table S3), risk classification (Table S4), and risk matrix (Table S5).

**Severity Scale:**
| Severity | Definition |
| --- | --- |
| Major | Resulting in serious injury to the patient and/or impedance to treatment |
| Moderate | Resulting in injury that will minimally impact patient care if at all |
| Minor | Resulting in no injury to the patient are considered expected flaws |

**Probability Scale:**
| Probability | Definition |
| --- | --- |
| High | Hazard will occur more often than not |
| Medium | Hazard will occur less often than it will |
| Low | Hazard rarely occurs |

**Risk Classification:**
| Risk Level | Definition |
| --- | --- |
| Level 1 | Unacceptable risk |
| Level 2 | Justifiable risk if limited by materials/cost limitations |
| Level 3 | Acceptable risk if negated by other methods utilized by healthcare workers (within reason) |
| Level 4 | No risk |

**Risk Matrix:**
|  | Severity |  |  |  |
| --- | --- | --- | --- | --- |
| Probability |  | Major | Moderate | Minor |
|  | High | Level 1 | Level 2 | Level 2 |
|  | Medium | Level 2 | Level 2 | Level 3 |
|  | Low | Level 3 | Level 4 | Level 4 |

**Table S6.**
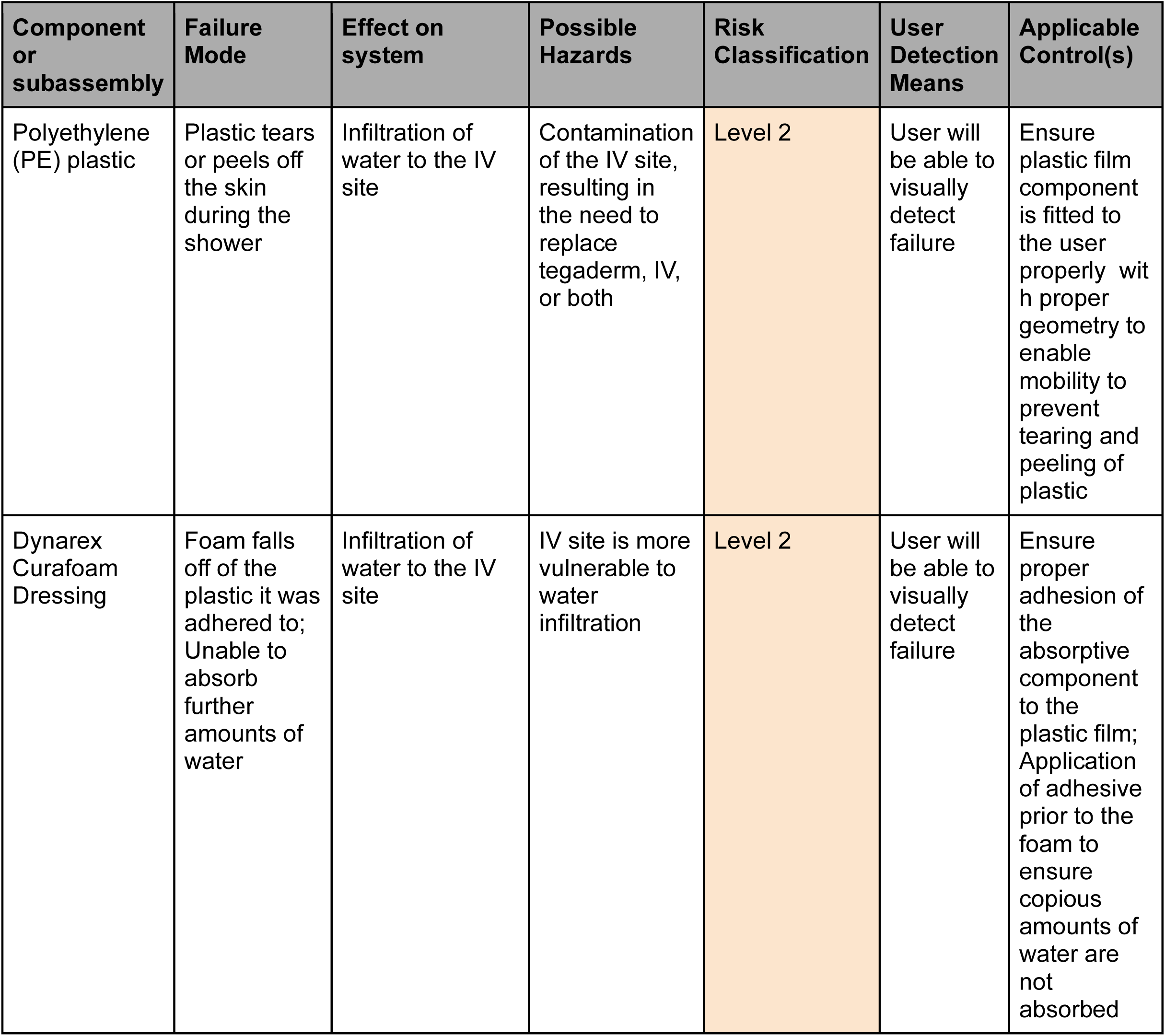
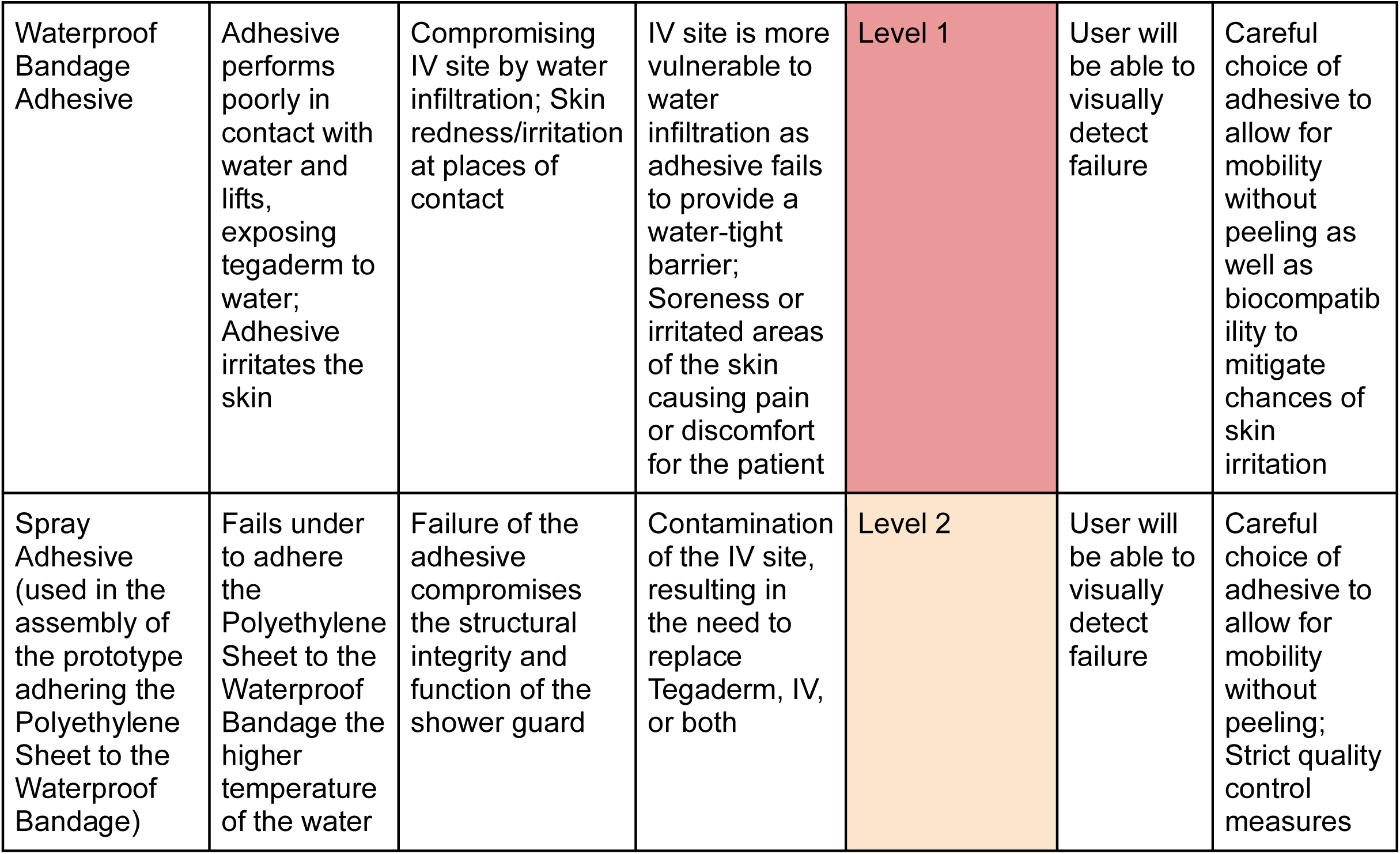
Table outlines potential hazards for each component involved in the construction of the DuckDerm. Tables S2-5 provide backing for the risk classification for each design failure mode that has been identified.

**Table S7.** Table outlines device specifications correlating to the qualitative user needs identified during ethnographic data collection. Functional, performance, and material requirements as well as use environment, are identified using the insights developed from the risk analyses. Key risks have been identified; design-based risk controls have been proposed along with residual risks introduced by the aforementioned design changes.

| Risk | Design-Based Risk Control | Residual Risks |
| --- | --- | --- |
| Water Intrusion | Choosing an appropriate waterproof bandage adhesive that will hold together the polyethylene plastic, foam padding, and the skin the product is applied to. | There is a reduced chance that water will infiltrate the IV site through failed adhesive. |
|  | Choosing a polyethylene plastic sheet with appropriate strength and flexibility to prevent tearing during use. | There is a reduced chance that water will infiltrate the IV site through tearing of the plastic. |
|  | Specifying design geometry to handle a wide range of motion, preventing tearing. This includes both the gap at the elbow for better range of motion without tearing as well as ensuring adhesive attaches the straps of the product to itself, creating a better seal. | There is a reduced chance that water will infiltrate the IV site through tearing of the plastic. |
|  | Choosing MedPride Foam Dressing material that can absorb large amounts of water. | There is a reduced chance that water will infiltrate the IV site through as there is a second barrier behind the adhesive to stop water from penetrating |
| Bacterial Contamination | Sterile packaging to prevent contamination before use. | Reduced risk of bacterial contamination prior to use. |
|  | Product is designed to be single-use and disposable to prevent buildup of contaminants between uses. | Product will not accumulate bacteria over time with each additional use. |
| Skin Irritation | Inclusion of directions/suggestions for the removal of the device. | If the user follows the directions, it will instruct them to remove with the direction of hair growth as to not irritate the skin. |
|  | Inclusion of directions and/or warnings for the proper application and use of the device so that it is applied correctly with the appropriate tightness. | User will be less likely to apply on incorrectly. |
| Dislodging of IV | Square of MedPride Foam padding placed around IV insertion site serves as stabilization for the dressing to dissuade dislodging due to excessive movement. | Patient will be less likely to disturb the inserted IV as it has a wall of protection around it |
|  | Inclusion of directions and/or warnings for the proper application and use of the device so that it is applied correctly with the appropriate tightness. | User will be less likely to apply on incorrectly. |

